# ‘It would nearly put the life back into you’ Older adults’ experiences of a Community Specialist Team for Older People (CSTOP) service model in Ireland: A Qualitative Descriptive Study

**DOI:** 10.1101/2024.12.13.24318927

**Authors:** Brian Condon, Anne Griffin, Collette Delvin, Christine Fitzgerald, Elaine Shanahan, Liam Glynn, Yves Couturier, Margaret O’Connor, Christina Hayes, Molly Manning, Rose Galvin, Aoife Leahy, Katie Robinson

## Abstract

**Introduction:** In Ireland, there has been a substantial recent investment in the Community Specialist Team for Older People (CST OP) service model. This approach provides timely integrated assessment and intervention for older adults in the community by a specialist multidisciplinary team. To inform the ongoing development and refinement of the CST OP service model, and ensure it is responsive to the needs and preferences of older adults, it is important to understand how older adults experience this new model of care. This qualitative descriptive study aims to resolve a research gap by exploring older adults’ experiences of the CST OP service model.

**Methods:** A qualitative descriptive study design was employed to explore older adults’ experiences of the CST OP service model. Purposive non-probability sampling was used to recruit 13 older adults who had completed intervention with a CST OP intervention. All interviews were completed in participants own homes, audio recorded and transcribed verbatim. A reflexive approach to thematic analysis guided data analysis.

**Findings:** Three themes were identified; older adults were uncertain about what to expect from the CST OP service and encountered accessibility barriers (theme1); the CST OP team provided coordinated, comprehensive care and built strong relationships with older adults (theme 2); CST OP intervention enabled older adults to better manage everyday activities and long-term conditions, thereby improving their wellbeing (theme 3).

**Discussion/ conclusion:** Our findings highlight the importance of CGA in community-based care for older adults. Further research is needed to address access barriers and evaluate older adults’ experiences with case management and care coordination in the CST OP service model.

## Introduction

Ireland currently has the fastest aging population in Western Europe (Sheehan and O’Sullivan, 2020). By 2051, it is projected that 26% of the population will be over the age of 65 (Sheehan and O’Sullivan, 2020). The World Health Organization (WHO) has identified that a fundamental shift in healthcare delivery is needed in response to population ageing, away from a fragmented care system, towards person-centred integrated care (WHO, 2015a).

Integrated care for older people refers to services that span the entire care continuum, integrating different levels and sites of care, while being tailored to meet individual’s needs (WHO, 2015b). Emerging research on integrated care models has demonstrated benefits for access to care (Baxter et al., 2018), quality of life, patient satisfaction, perceived quality of care (Baxter et al., 2018; Uittenbroek et al., 2016; Hayes et al., in press; Yan Wang et al., 2024) and weak beneficial effects on health outcomes for older adults (Kirvalidze et al., 2024). Additionally, these co-ordinated community-based approaches may reduce hospital admission rates and lengths of hospital stay for older adults (Liljas et al., 2019).

In Ireland, the Integrated Care Programme for Older Persons (ICOPOP) aims to develop and implement integrated services and pathways for older people with the development of community based, planned and coordinated care (ICPOP, 2017). A key component of this program is the Community Specialist Team for Older People (CST OP) service model of care. CST OP teams typically comprise a range of community-based healthcare professionals, including nurses, physiotherapists, occupational therapists, case manager, speech and language therapist’s, dietitians and consultant geriatricians (Health Service Executive, 2024a). These teams enable timely access to Comprehensive Geriatric Assessment (CGA) where the different healthcare professionals work together to assess and to respond to an older person’s medical, functional and social needs (Health Service Executive, 2024a). In this model of care primary and secondary care services work together to avoid unnecessary hospital admissions for older adults (Fitzgearld et al., 2023).

The British Geriatrics Society has advocated for the involvement of older adults in the co-design, delivery, and ongoing monitoring of Integrated Care services (British Geriatrics Society, 2024), a view echoed in Irish integrated care policies (ICPOP, 2017) and in academic literature (Sadler et al., 2019). Research suggests that there may be substantive differences between healthcare providers perspectives of integrated care and older adults’ perspectives (Sheaff et al., 2017). Through their qualitative systematic review, Karascony and colleagues identified a lack of older adult voices, particularly from the oldest-old population, in the current evidence base on the clinical and social outcomes of integrated care (Karascony et al., 2022). This suggests that further research is needed to more comprehensively capture the experiences and needs of this demographic, who may have distinct requirements and preferences compared to younger older adults.

## Methodology

### Study Aim

The aim of this research is to explore older adults’ experiences of the CST OP service model within the Irish Healthcare context.

### Design

A qualitive exploratory descriptive approach was adopted (Daly et al., 2007) using semi-structured interview methodology and reflexive thematic analysis (Bruan and Clarke, 2019) to explore older adults’ experiences of CST OP model of care in the Mid-West region of Ireland. Given that the CST OP service model was newly formed, this research design aligned with our aim of presenting comprehensive detailed summary of participants experiences and without use of a theoretical framework (Kim et al., 2017). This study was conducted in parallel to a cohort study exploring older adults’ health outcomes following intervention by CST OP (Hayes et al., in press). The conduct and reporting of this study were in accordance with the Consolidated Criteria for Reporting Qualitative Research (COREQ) checklist (supplementary file 1) for focus groups and interviews to ensure rigor, comprehensiveness, and credibility of the research study (Tong et al., 2007).

### Context / setting

The setting of participant recruitment was from three CST OP services within Ambulatory Care Specialist Hub (ACSH) sites (Ennis, Limerick and Thurles) in the Mid-West region of Ireland which has a population of just over 400,000 (Health Service Executive, 2024b). The profile of the population in this region is older and more deprived than the national comparator (Health Service Executive,2024b). Additionally, between 2016 and 2022, the 75–79-year age group increased by almost 40%, and the 85 years and older age group increased by 25% in this region (Health Service Executive,2024b). Older adults were recruited after their intervention with CST OP team thus, inclusion and exclusion criteria for prospective participants to the study was based on referral criteria to the CST OP team as outlined in the table 1.1

**Table 1.1.**
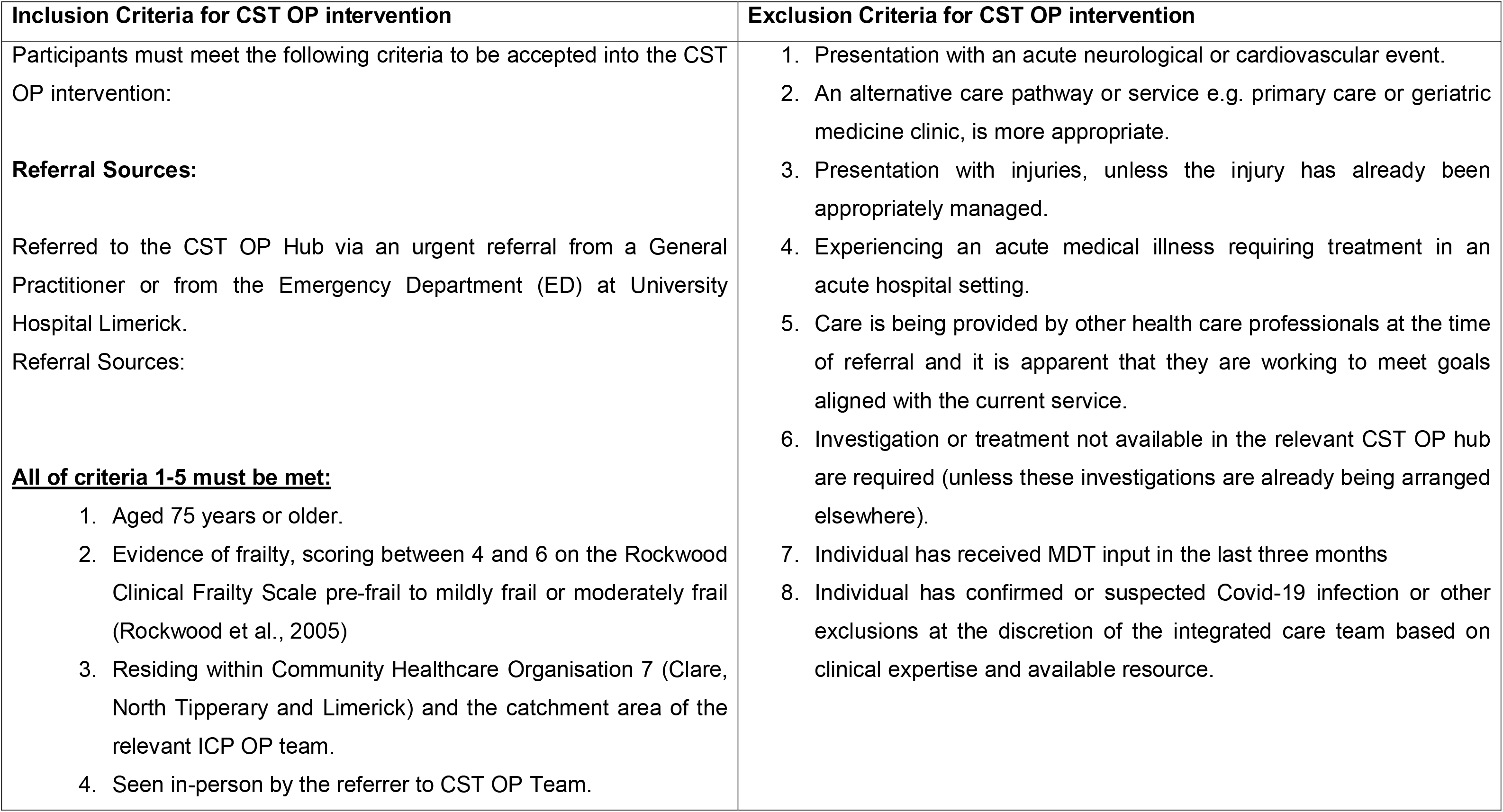
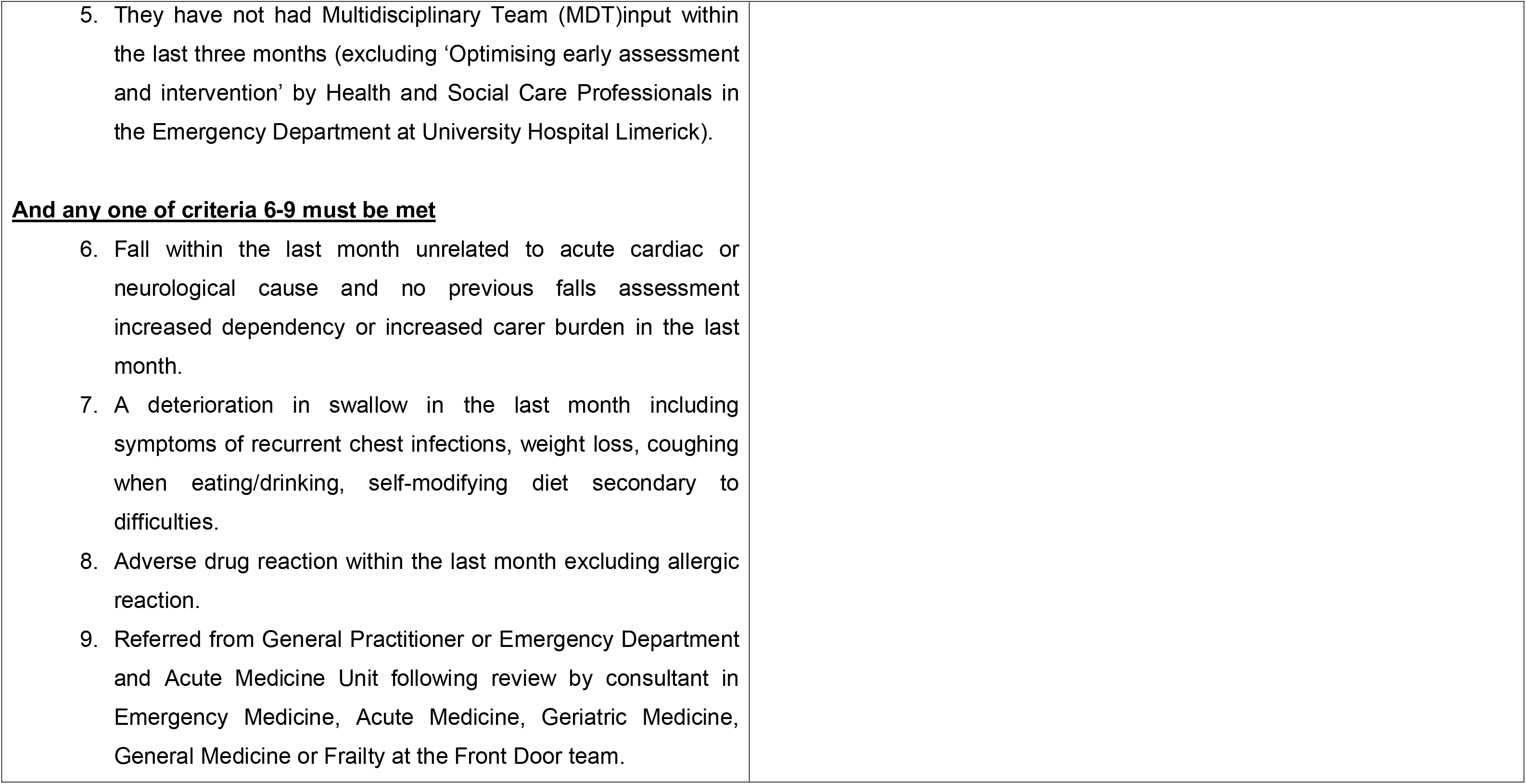
Inclusion and exclusion criteria.

### Sampling and recruitment

Purposive non-probability sampling was applied to select prospectively older adult participants who had completed intervention with the CST OP. A research nurse (CD) involved in the parallel cohort study acted as a gatekeeper by providing study information (participant information leaflet, consent form) to potential participants who met the inclusion criteria. Alongside inclusion criteria, the gatekeeper considered variation of participants regarding gender, age, and social circumstances (living alone /with others) as participant recruitment progressed. Participants were equally recruited from the three CST OP sites (rural and urban settings). After explaining the study aim and providing an opportunity to ask questions, if a prospective participant expressed an interest in taking part, the research nurse (CD) obtained consent to share their contact details with the male qualitative interviewer (BC). The research nurse (CD) and interviewer (BC) had no direct care involvement with prospective participants. The interviewer (BC) contacted them by phone and an interview date/time was scheduled.

### Positionality & reflexivity

Participant interviews, interview transcription, open coding of transcripts and data analysis were conducted by the lead author (BC) under supervision of the wider research team. The lead author is a registered occupational therapist with over fourteen years clinical experience working with older adults and is a PhD candidate who has completed postgraduate training in qualitative research. Interviewees knew he was a PhD candidate and did not know he was an occupational therapist. Members of the wider research team have clinical and qualitative research experience with older adults in their respective roles: general practitioner (LG), geriatrician (MOC, ES, AL), physiotherapist (RG, CH), dietitian (AG), speech and language therapist (MM), nurse (CF, CD) and occupational therapist (KR, BC).

### Data Collection

All interviews were conducted in participants homes at various locations in the Mid- West region and each interview was conducted by the same interviewer (BC). On occasion, at the request of participants (n=4), family members were present during the interviews. This was especially helpful for participants with sensory impairments, as family members could clarify if older adults had heard the interview questions posed by the interviewer (BC). However, family members did not answer on participants behalf and no data collected from participant family members was used within this study. No repeat interviews were required.

At the outset of each interview, participants were given an opportunity to ask any questions about participating in the research study. Those who agreed to participate signed two copies of a consent form (one was kept by the researcher, and one remained in the participants home). Field notes were taken after each interview and recruitment ceased when data saturation was reached, the point at which there was no further additional emergence of issues/ views with repetition of same data thus, making further data collection redundant (Hennick et al., 2022). Saturation was determined by the interviewer (BC) and another member of the research team (KR)

Individual semi-structured interviews were conducted with the interview topic guide developed from findings from a recent study exploring older adult’s experiences transitioning from the ED (Condon et al., 2024) and a literature review on the topic area. Additionally, an older person and family caregiver Public and Patient Involvement Panel, and research experts in the area were consulted. The topic guide focused on three main concepts: overall experience at the CST OP, experience of care delivery at the CST OP, experience of communication and the discharge process at the CST OP (see supplementary file 2). A pilot of the semi structured interview guide was completed by the lead researcher (BC) with an older adult who provided feedback on length and focus of the topic guide questions, no amendments were made following the pilot. The pilot interview was included in the study. The semi structured interviews were audio-recorded, transcribed in full, and exported to NVivo version 14 Pro Software (Lumivero,2024). Transcripts were not returned to participants for comment and correction to ensure integrity of the data as all audio recordings were clear and high quality for accurate transcription (Morse et al., 2015).

### Data Analysis

A reflexive approach to thematic analysis as described by Braun and Clarke guided the analysis of the data (Bruan and Clarke, 2019). This reflexive approach aligns with the qualitative descriptive design and is appropriate given the lead researchers dual role as both a researcher and a clinician (Braun and Clarke, 2019). The emphasis on researcher reflexivity in this approach supported identification of the researcher role at all stages of the study. An iterative approach to analysis was applied to which data collection and analysis occurred concurrently to inform one another (Morgan and Nica, 2020). Data was analysed inductively to generate analytical codes using a ‘bottom up’ approach, rather than deductive, ‘top-down’ application of a priori theories or frameworks. Themes were identified on a semantic level (explicit or overt meanings), placing the focus explicitly on transcript content (Braun and Clarke, 2019).

The six steps outlined by Braun and Clarke were conducted: familiarisation with data, generating initial codes, searching for themes, defining, and naming themes and producing the report were followed in the data analysis stage (Bruan and Clarke, 2019). In step one, the lead author (BC) read and re-read the transcripts in detail in a process of in-depth data immersion and familiarisation. This was followed by the lead author generating initial codes (forming a coding tee) using NVivo version 14 Pro Software (Lumivero,2024). The lead author had attended workshops on thematic analysis. Concurrently, the coder (BC) gained consensus on major topics and subtopics with two members of the research team (KR, AG) who have established qualitative expertise. This step was achieved through frequent research meetings involving critical discussion and debriefing between three research members (BC, KR, AG).

The third step of thematic analysis ‘searching for themes’ was commenced by three members of the research team (BC, KR, AG) in identifying overlapping codes leading to generated themes representing a pattern of data from the interviews (Braun and Clarke, 2006). Step four involved reviewing potential themes by members of the research team (BC, KR, AG). This involved judging whether there was sufficient interview data to underpin each generated theme. In step 5 ‘naming the themes’ clear names and definitions were applied to each generated theme. In the final step a final report in the version of this manuscript was completed. The findings of the study were not shared with participants as further research was planned in this research area.

### Ethical considerations

Ethical approval was gained from the Health Service Executive (HSE) Mid-Western Regional Hospital Ethics Committee at University Hospital Limerick in December 2021 (Research Ethics Committee Reference:116/2021). Interviews only commenced when informed consent from participants was received and to protect their privacy and confidentiality, all participants names were de-identified with each participant provided with a unique participant number. The gatekeeper (CD) and lead researcher (BC) were the only members of the research team who could identify the data. Any identifiable information (e.g. patients name, home address etc.) was removed or adapted from participants quotes used to illustrate findings within this study.

### Participant Characteristics

Sixteen older adults were initially recruited, two participants did not return a consent form and subsequently were not interviewed, and one participant withdrew from the study after being interviewed. All thirteen interviews included in this study were conducted between April 2023 and September 2023, with interviews lasting between 20 and 60 minutes. Data from interviews with thirteen older adults, female (n=10) and male (n=3) are presented. Participants reported diverse socio-economic backgrounds based on Pobal Haase and Pratschke Deprivation Indices (Haase and Pratskhke, 2022): marginally above socio- economic (n=7), marginally below socio- economic (n=5) and disadvantaged socio-economic (n=1). Additionally, participants were recruited/included from both rural (n=2) and urban settings (n=11). Participants demographics is detailed in table 1.2

**Table 1.2.** Participant demographics. MT=Medical Team, PT= Physiotherapy, OT= Occupational Therapy, Diet= Dietician, CM= Case Manager, SLT= Speech and language therapist, ANP= Advanced Nurse Practitioner, CNS=Clinical Nurse cialist

| **Participant Number** | **Gender** | **Length of recording** | **Age** | **Ethnicity** | **Residential Status** | **Presenting problem** | **Disciplines seen at CST OP** |
| --- | --- | --- | --- | --- | --- | --- | --- |
| 1 | F | 33.49 mins | 75-80 | White Irish | Lives alone | Falls | MT, PT, Diet. |
| 2 | M | 53.49 mins | 85-90 | White Irish | Lives alone | Falls | MT, PT, OT |
| 3 | F | 20.55 mins | 80-84 | White Irish | Lives with family | Reduced mobility | MT |
| 4 | F | 20.55 mins | 80-84 | White Irish | Lives with family | Falls | MT |
| 5 | F | 38.29 mins | 75-80 | White Irish | Lives with other | Parkinson disease. | MT, PT, SLT, CNS |
| 6 | F | 26.03 mins | 90-95 | White Irish | Lives with other | Dizziness | MT, PT, CNS |
| 7 | M | 30.55 mins | 90-95 | White Irish | Lives with other | Falls | MT, ANP, PT, SLT |
| 8 | F | 31.01 mins | 90-95 | White Irish | Lives alone | Falls | MT, CNS, PT, |
| 9 | F | 32.11 mins | 80-84 | White Irish | Lives alone | Falls | MT, ANP, PT |
| 10 | F | 31.11 mins | 90-95 | White Irish | Lives alone | Functional decline | MT, ANP, CM, PT, SLT, OT |
| 11 | F | 13.18 mins | 90-95 | White Irish | Lives with other | Falls | MT, CNS, CM, SLT |
| 12 | F | 28.08mins | 75-80 | White Irish | Lives with other | Increased Frailty | MT, CNS |
| 13 | F | 58.36 mins | 75-80 | White Irish | Lives with other | Falls | MT, CM, CNS, PT |

Participants ages ranged from 77 to 95 with a mean age of 85.38 years of age. The presenting problem for participants attending CST OP included falls (n=7), dizziness (n=1), increased frailty (n=1), Parkinson disease (n=1), and functional decline (n=2). All included participants were seen by a medical team member of the CST OP team (n=13) with only four participants not seen by the physiotherapist (n=11) and four not seen by nursing staff (n=12).

### Findings

Three overarching themes were generated from the data which will be presented as our findings. Older adults were uncertain about what to expect from the CST OP service and encountered accessibility barriers (theme 1). The CST OP team delivered positive, coordinated, and comprehensive care, and developed strong therapeutic relationships with older adults (theme 2). CST OP intervention enables older adults to better manage everyday activities and long-term conditions, enhancing emotional well-being (theme 3).

**Theme 1: Older adults were uncertain about what to expect from the CST OP service and encountered accessibility barriers.**

This theme relates to older adults’ expectations of CST OP prior to engaging with the service. They welcomed being referred to the service however, many reported a lack of prior knowledge about the service prior to attending, leaving them uncertain about what to expect. Furthermore, some older adults described difficulties attending appointments due to due to transportation dependencies.

Prior to attending the CST OP, symptoms and challenges reported by older adults included falls, pain, infections, palpitations, reduced confidence, social isolation, and grief. In certain situations, these symptoms impacted on their ability to manage at home for example, one participant described how they were afraid to go up and down the stairs at home. Additionally, another participant described how their palpitations were disturbing their sleep.

**‘I was on crutches for a long time and my shoulders then were giving me a lot of problems, so I had physical and mental problems. As a result, you know the grief I went through and then to find I couldn’t walk’ (Participant 9)**

Prior to CST OP some participants required spousal, family or formal caregiver assistance to complete daily activities. By attending CST OP, older adults hoped to regain lost abilities, such as return to driving, increased walking, and stair climbing. Older adults interviewed in this study reported that, both the General Practitioners (GPs) and hospital doctor who referred them to the CST OP service had limited knowledge about the service. Additionally, several participants reported that they were unaware of who and why they were referred to CST OP.

**‘I don’t know which GP that referred me to the HUB. But I am thrilled that they did and for what reason, I don’t know. I don’t know why they sent me there now it could be because of my back’ (Participant 13)**

Most participants indicated that their referral to CST OP was probably catalysed by their reduced function, need to access physiotherapy, or frailty.

Older adults reported receiving varying levels of information about CST OP prior to attending the service, ranging from verbal explanations from referrers to appointment letters from CST OP, but participants generally described limited understanding from their GPs or hospital doctors about CST OP’s role in their care. A common description provided to older adults by their GP was that CST OP was a ‘new service’.

**‘Yes (GP), he explained it all that it was a new, what would you say, a new program that they were starting up and that he asked me whether if I would like to do it firstly’ (Participant 1).**

Some older adults described accessibility challenges in attending CST OP, including difficulties arranging transportation as they were reliant on others for transportation and issues with parking at CST OP location.

**‘I never learned to drive so I don’t drive so I’ve always have to get a friend to drive me over or my son would drive me over’ (Participant 7).**

Some participants described that, they had to travel outside their local community to an urban setting to attend CST OP. One older adult described frustration that geographical boundaries limited them attending their nearest CST OP team.

**Theme 2: The CST OP team provided coordinated, comprehensive care and built strong relationships with older adults.**

This theme relates to older adults’ experiences of the CST OP team. They described thorough assessments and quick access to medical testing, leading to timely diagnoses when attending CST OP. This promptness of assessments and medical testing was followed by well-planned, comprehensive care, and the strong relationships formed with CST OP team members enhanced their overall experience with the service.

**’He (GP) couldn’t believe all that was checked out, with my like my blood work, and everything was checked out and he got all the results, and they were all within normal range and thank God I don’t have high blood pressure or low blood pressure its quite normal’’ (Participant 12).**

Older adults described that, they were impressed by the rapid access to diagnostics including bone density scans, blood tests, head up tilt tests, memory assessments and 24hour blood pressure monitors. These services and medical diagnostics facilitated prompt diagnosis and effective management of their health complaints.

**‘’And what I was very impressed was immediately when I told them about the hip and the fracture, they send me for a DEXA (dual energy X-ray absorptiometry) scan which showed I had osteoporosis’’ (Participant 9).**

In some cases, when results of these diagnostic test results were normal, it reassured older adults or enhanced their confidence with managing their condition or symptoms at home independently.

**‘Well naturally when you go over 80 not to mention going over 90, you’re nervous … when you get good news its very encouraging to keep going and to fight and to keep, keep fighting and don’t sit down and give in’ (Participant 8)**

Older adults reported that the assessments and examinations conducted by the CST OP team was very thorough. They felt that every aspect of their healthcare concerns and needs was addressed in detail.

**‘I got an overhaul from head to toe. They didn’t leave anything out else they examined me every way … I found it very good now’ (Participant 9).**

In contrast to other healthcare experiences older adults described that, there was no time limits imposed when voicing their health concerns to the CST OP team. They appreciated the opportunity to fully explain their issues without the feeling of being rushed. The comprehensive nature of the assessment, along with the open dialogue with the CST OP team, allowed the older adults to feel heard and understood in a way that was distinct from their typical healthcare encounters.

**‘Oh yeah, you could tell them anything. You could tell them anything there was no problem there’ (Participant 7).**

The positive communication with the CST OP team resulted in, older adults feeling relaxed and comfortable which enabled them to share important information and priorities.

**‘I don’t know you’re just feel comfortable you know, I felt comfortable in my own skin whereas you go down to the doctor you get tensed up before you go in’ (Participant 13).**

Older adult’s descriptions of the CST OP team members and the relationships they established with the team were universally positive. CST OP team members were described as kind, generous, caring, nice, helpful, understanding, empathic and concerned with older adults, leading to a comfortable assessment and intervention process for older adults.

**‘It was their kindness and understanding and you know you can sit here like, make sure I could sit down and was able to walk on my own and if not, they were going to get a wheelchair’ (Participant 9).**

**Theme 3: CST OP intervention enabled older adults to better manage everyday activities and long-term conditions, thereby improving their wellbeing.**

This theme related to older adults’ description that intervention from CST OP enabled them to better manage their long-term conditions and improved their ability to complete activities and maintain independence at home. This ultimately led to improved older adult emotional wellbeing.

Older adults described presenting to CST OP with a range of long-term conditions and symptoms and concerns about their health that, impacted their ability to manage at home. Older adults described how intervention and rehabilitation from CST OP enabled them to better manage their long-term conditions and improved their ability to complete activities and maintain independence at home.

**‘Yes, they did help me in in that way, like being far more confident, you know, going up and down the stairs’ (Participant 9)**

Older adults described that a medication review as a key component of the CST OP intervention. Participants describe that in comparison to other healthcare services they previously had accessed, detailed consideration was taken by CST OP of their medication regime. They described various changes to their medications following this review which they attributed to various positive outcomes such as improved symptom management (e.g. pain, high / low blood pressure and infections) and better management of chronic conditions. For example, one participant described following a change to their medication regime by CST OP it enabled effective management of their palpitations leading to their increased independence at home.

**‘They put me on the tablet you know what I mean and the tablet slowed down the palpitations so that I am very thankful for because, I mean I don’t know where I’d be now if because, it was nearly coming up my throat the palpitations were going, the heart would take off if I walk from here and, I get breathless if I walk to the sink and they found that so that I’m thankful for it’ (Participant 13)**

Older adults valued home visits (a small number of older adults had home visits by an OT) and provision of assistive devices, strategies, and exercise plans by the CST OP team to manage long term conditions and maintain/ improve safety and independence in daily activities.

**‘I did admit that it was difficult using the bath, which I use every night or most nights. And I got the chair for the bath, they installed that for me, and I find that is a marvellous help’ (Participant 10)**

Older adults reported receiving clear, detailed exercise plans tailored to their individual needs, with thorough explanations from the CST OP physiotherapist. Older adults greatly valued these personalised exercise plans which enabled them to continue the exercise plans on their own after discharged from the service, and ultimately enabled them to maintain their mobility and independence. Other reported benefits of exercise programs included improved quality of life and reduced pain.

**‘I found that I kept doing my exercises and that I am walking an awful lot better, and my quality of life has improved’ (Participant 12)**

One participant described how they were able to achieve their rehabilitation goal of being able to walk independently with a walking stick after their intervention with the CST OP service.

**‘Well as I said they got me back walking again properly, because I couldn’t walk before, I’m alright now walking … She had me walking round first of all in a frame and then at the end of it, she had me onto a stick’ (Participant 6)**

One participant highlighted specific health benefits of CST OP intervention, noting how exercises from the CST OP Speech and Language Therapist effectively addressed their swallowing issue.

**‘She was telling me how to chew my food and that type of thing well…..when I was eating at times, I’d get a cough you know, and she dealt with that and dealt with it very capably’ (Participant 7)**

Older adults described positive psychological, well-being changes along with improved quality of life following their intervention with CST OP. In many cases, improvements in their function led to improved emotional well-being. Within this study, older adults provided a multitude of examples of returning to their usual life activities/hobbies and increased independence/improved performance in these activities due to the intervention from the CST OP team. One participant described intervention from CST OP as transformative.

**‘It was like, it was like you know it (CST OP) would nearly put back the life back into you’ (Participant 13)**

Older adults described new feelings of confidence and liberation after their intervention with the CST OP team. Older adults gave examples that intervention from CST OP enabled them to return to activities that, they had stopped prior to CST OP such as driving to visit their family members or being able to do their stairs at home.

**‘But shortly after the, my final visit, I drove to Dublin, you know, and I found that that was very liberating. Actually, I can do it again now you know’ (Participant 9)**

## Discussion

### Summary of main findings

This qualitative descriptive study aimed to explore older adults’ experiences of the CST OP service in the Mid-West of Ireland. From interviews with 13 participants attending the service we found that, older adults highly valued the service. However, many faced accessibility barriers and were unsure of what to expect from the service prior to participating (theme 1). Older adults perceived that the CST OP team delivered coordinated, and comprehensive care, developing strong relationships with them (theme 2). Importantly, the CST OP intervention was empowering for the older adults, enabling them to better manage their long-term conditions and maintain their independence living at home. Overall, the CST OP service was experienced by older adults to have a positive impact on their functional ability and emotional well-being (theme 3).

### Comparison with existing literature

Our findings offer a comprehensive account of older adults’ experiences with a new model of community-based, integrated care for older adults in Ireland. This demographic is often underrepresented within clinical research, highlighting a crucial need for health research that includes this population cohort (Thake et al., 2017). Notably, previous qualitative research on integrated care experiences has largely overlooked the perspectives of older adults (Karascony et al., 2022). Within healthcare research, older adults aged 85 years and older are often referred to as the ‘oldest old’ (Gu & Dupre, 2021) and research on preventative interventions for this population is limited (Escourrou et al., 2022). Within this study, participants ranged in age from 76 to 95 years with a mean age of 85 years. Our findings provide new knowledge for decision makers and policy makers on the integrated care experiences and perceptions of an ‘oldest old’ demographic cohort.

Most participants within this study required assistance from family or others for transportation to CST OP sites and therefore faced accessibility issues in attending the service. This finding is keeping with data from the Irish Longitudinal Study on Ageing (TILDA) which revealed that, as older adults age in Ireland, they are less likely to drive and become reliant on others for transport due to poor public transportation (Donoghue et al., 2017). The WHO has recommended that integrated care services should be offered close to where people live (WHO, 2018). Additionally, recent qualitative systematic review found that older adult’s express greater comfort and confidence when they can receive integrated care services within their own homes (Karascony et al., 2022). Provision of transport or provision of home-based services are two potential solutions both with potential cost and acceptability implications. Integrated care has been found to likely reduce cost however, the evidence varies largely and is only of moderate quality (Rocks et al., 2020). A review of cost-effectiveness of homecare services compared to in-hospital care for adults concluded that, homecare interventions are likely to be cost-saving and as effective as care in a hospital (Curioni et al., 2023). However, little is known about how well public transport systems provide older adults with access to key destinations in various regions around the world (Ravensbergen et al., 2022) thus, further research to explore transport interventions and provision of CST OP services in the home environment are warranted.

A primary finding from this study was that older adults valued the comprehensive CST OP assessment. CGA has been extensively researched and demonstrated to yield quantifiable positive healthcare outcomes for older adults across a variety of settings, including primary care (Pilotto et al., 2017; Garrard et al., 2020; Welsh et al., 2014). The use of CGA within integrated care services for older adults is also actively promoted by the WHO (WHO, 2018). Our findings indicate that older adults valued CGA as it effectively addressed their healthcare concerns, managed their presenting symptoms, and facilitated timely access to necessary interventions and therapies. These findings align with those of a recent qualitative evidence synthesis, which reported that older adults who underwent a CGA in community-based settings experience it positively as it promotes a holistic approach in addressing their needs (Hayes et al., 2023).

Medication reviews are one of the most common elements of integrated care models for older adults (Briggs et al, 2018) and unsurprisingly are core to the CST OP model. We found older adults experienced medication review as leading to significant adjustments and positive outcomes including improved symptom management. A recent cohort study by Payen and colleagues found that combining a medication review with an integrated care approach was linked to a reduction in the number of hospital readmissions within 30 days for older adults (Payen et al., 2022). Given that polypharmacy is common among older adults due to multiple co-mobilities this group are at risk of inappropriate prescriptions or adverse drug effects (Beuscart et al., 2021). Medication safety has been identified as a key area for improvement for all healthcare settings in line with WHO recommendations (HIQA, 2019).

In contrast with the broader literature on integrated care, findings do not provide rich information on older adult’s experiences of care-coordination or communication between professionals, their involvement in care planning and decision-making, or if there were any gaps or fragmentation in providers/services that disrupted their care experience. This may reflect the differing priorities of heath care between older adults and healthcare researchers/healthcare providers.

A case management approach is being embedded with CST OP services (Barry et al., 2021). Case management is a role undertaken by a health or social care professional, supported by a wider team with responsibilities including assessment, care planning, and co-ordination of care to meet the needs the older adult (Sadler et al., 2023). Within this study, older adults described meeting the same nurse or professional from the CST OP during their intervention which may have been an healthcare professional working within a case manager role. Of note, a recent Cochrane review examining case management programmes for older people living with frailty in the community found they may make little or no difference to patient and service outcomes and care-related costs (Sadler et al., 2023). Therefore, further research is required on the role and effectiveness of case management within CST OP model of care.

Communication with healthcare providers in integrated care service is of critical importance as older adults typically define their perspectives towards integrated care with respect to the relational, informational and organisational aspects of care (Lawless et al., 2020). We found that older adults had positive experiences communicating with CST OP teams, such as feeling unhurried, feeling comfortable sharing information, and receiving clear information/ instructions from team members. This is important as the quality of healthcare provider communication with older adults can impact their trust and satisfaction with their healthcare provider, impacting overall health and outcomes for older adults (Birkhäuer et al., 2017). Additionally, older adults are more likely to talk about their healthcare problems when healthcare professionals employ an open style of communication and listen to their opinions in relation to their care (Gibney & Moore, 2018) which aligns with the process of CGA which underpins the CST OP model of care.

### Strengths and Limitations

This is the first study to explore older adults’ experiences of the CST OP service model in Ireland. We acknowledge the lack of participant diversity, as all recruited participants identified as ‘White Irish’. However, our participant profile sample is reflective of the 2022 national Census which identified that, most older adults aged 85 years and older identify their ethnicity as being White Irish (CSO, 2024). Ease and comfort throughout interview process of participants was maximised by conducting interviews in participants’ homes and having caregivers present, when requested by participants. However, it is possible that the presence of caregivers may have influenced participant responses to interviewer questions. Additionally, some older adults that had intervention with CST OP were also accessing multiple healthcare services, thus had difficulty in distinguishing their intervention with CST OP from other healthcare encounters.

This research study has a sample size of 13 and data saturation was achieved. The qualitative analysis was enriched by the researchers prolonged engagement with the data, rigorous approach to coding and adherence to principles of investigator triangulation during analysis. The concept of this study was informed by an established Public and Patient Involvement (PPI) stakeholder panel of older adults and family caregivers. Additionality, semi-structured interviews were underpinned by rigorous literature review and was piloted with an older adult to develop and refine topic guide. A further strength of the study was that the research team were from different healthcare backgrounds, with diverse clinical and research experience.

## Conclusion

This study offers in-depth qualitative exploration of older adults’ experience of the CST OP service in the Midwest region of Ireland. Our findings indicate that older adults highly valued the CST OP service, however, were uncertain of what to expect prior to engaging with the service and faced some accessibility barriers attending the hubs (Theme 1); the CST OP team delivered coordinated, comprehensive care and fostered strong relationships (Theme 2); and the CST OP intervention enabled older adults manage daily activities and long-term conditions, enhancing their well-being (Theme 3).

Our findings underscore the value of CGA in community-based care to meet the needs of older adults. Further research is needed on solutions to access barriers, as well as further evaluation of older adults’ experiences with case management and care coordination within the CST OP service model.

## Data Availability

All relevant data are within the manuscript and its Supporting Information files.

## Abbreviation List

ICPOP: Integrated Care Programme for older persons
CST: Community Specialist team
CGA: Comprehensive Geriatric Assessment
WHO: World Health Organisation
ADL: Activities of Daily Living
MDT: Multidisciplinary team
ACSH: Ambulatory Care Specialist Hub
GP: General Practitioner
PT: Physiotherapy
OT: Occupational Therapy
ANP: Advanced Nurse Practitioner
SLT: Speech and Language Therapy
CNS: Clinical Nurse Specialist
CM: Case Manager MT Medical Team
ED: Emergency Department
HSE: Health Service Executive
MDT: Muli-Disciplinary Team Members
DEXA: Dual energy X-ray absorptiometry

## Ethics approval and consent to participate

Ethical approval was gained from the HSE Mid-Western Regional Hospital Ethics Committee at the University Hospital of Limerick (Rec Ref:116/2021).

## Availability of data and materials

The datasets from this research study are not readily available to protect participants confidentiality. Requests to access anonymised partial datasets can be made by contact the lead author.

## Authors’ contribution

BC was responsible for conceptualisation of the study, design of the work, analysis, and interpretation of the data, and writing the initial draft of the manuscript. KR, AG, RG, MM and CF were responsible for conceptualisation of the study, design of the work, analysis and interpretation of the data, supervision, and revised the manuscript from a critical perspective to enhance its contents. All the authors have read and approved the final draft of the manuscript.

## Consent for publication

Not applicable

## Acknowledgements

The research team would like to acknowledge the participants recruited in this study for their time given to complete this study.

## Supplementary File 1

### COREQ Checklist

| Topic | Item No | Guide Questions/description | Reported on page no |
| --- | --- | --- | --- |
| **Domain 1: Research team and reflexivity** |  |  |  |
| Personal Characteristics |  |  |  |
| Interviewer/facilitator | 1 | Which author/s conducted the interview or focus group? | 8 |
| Credentials | 2 | What are the researcher credentials? E.g. PhD,MD | 8 |
| Occupation | 3 | What was their occupation at the point of their study? | 8 |
| Gender | 4 | Was the researcher male or female? | 8 |
| Experience and training | 5 | What experience or training did the researcher have? | 8,10 |
| Relationship with participants |  |  |  |
| Relationship established | 6 | Was a relationship established prior to study commencement? | 8,9 |
| Participant knowledge of the interviewer | 7 | What did the participants know about the researcher? e.g. personal goals, reasons for doing the research. | 8,9 |
| Interviewer characteristics | 8 | What characteristics were reported about the interviewer/facilitator? e.g. Bias, assumptions, reasons and interests in the research topic. | 8 |
| **Domain 2: Study Design** |  |  |  |
| Theoretical framework |  |  |  |
| Methodological orientation and Theory | 9 | What methodological orientation was stated to underpin the study? e.g. grounded theory, discourse analysis, discourse analysis, ethnography, phenomenology, content analysis | 4 |
| Participant selection |  |  |  |
| Sampling | 10 | How were the participants selected? e.g. purposive, convenience, consecutive, snowball | 8 |
| Method of approach | 11 | How were participants approached? e.g. face-to-face, telephone, mail. email | 8,9 |
| Sample size | 12 | How many participants were in the study? | 11 |
| Non-participation | 13 | How many people refused to participate or dropped out? Reasons? | 11 |
| Setting |  |  |  |
| Setting of data collection | 14 | Where was the data collected? e.g. home, clinic, workplace | 8 |
| Presence of non-participants | 15 | Was anyone else present besides the participants and researchers? | 8 |
| Description of sample | 16 | What are the important characteristics of the sample? e.g. demographic data, date. | 12 |
| Data collection |  |  |  |
| Interview guide | 17 | Were questions, prompts, guides provided by the authors? Was it pilot tested? | Supplementary File 2 |
| Repeat interviews | 18 | Were repeat interviews carried out? If yes, how many? | 9 |
| Audio/visual recording | 19 | Did the research use audio or visual recording to collect the data? | 9 |
| Field notes | 20 | Were field notes made during and/or after the interviewer or focus group? | 9 |
| Duration | 21 | What was the duration of interviews of focus groups? | 12 |
| Data saturation | 22 | Was data saturation discussed? | 9 |
| Transcripts | 23 | Were transcripts returned to participants for comment and/or correction? | 9 |
| **Domain 3: analysis and findings** |  |  |  |
| Data analysis |  |  |  |
| Number of data coders? | 24 | How many data coders coded the data? | 10 |
| Description of the coding tree | 25 | Did authors provide a description of the coding tree? | 10 |
| Derivation of themes | 26 | Were themes identified in advance or derived from the data? | 9,10 |
| Software | 27 | What software, if applicable, was used to manage the data? | 10 |
| Participant checking | 28 | Did participants provide feedback on the findings? | 10 |
| Reporting |  |  |  |
| Quotations presented | 29 | Were participants quotations presented to illustrate the themes/findings?<br>Was each quotation identified? e.g. participant number | 13-18 |
| Data and findings consistent | 30 | Was there consistency between the data presented and the findings? | 13-18 |
| Clarity of major themes | 31 | Were major themes clearly presented in the findings? | 13-18 |
| Clarity of minor themes | 32 | Is there a description of diverse cases or discussion of minor themes? | 13-18 |

## Supplementary File 2

### Topic Guide

#### Main Theme: At the Ambulatory Care Hub

1. Why did you initially attend the Ambulatory Care Hub (ACH)?
2. What did you know about the team or the service before you went? Did you know why you were there? (Understand the role of the ACH in your care vs for e.g., GP care)
3. What was your experience in attending the ACH? Could you describe it for me? (Travel, transport, waiting room, space, meals, length of time, parking).
4. Who did you see in the ACH? (Different team members)
5. Did each team member explain their role in your care?
6. How many visits did you have to the ACH?

#### Main Theme: Delivery of care

1. Could you describe for me what your experience of the care/intervention/therapy delivered by the ACH?
2. Did you have a say in your treatment plan? (Goal – setting)
3. During your sessions with the team, did anybody talk how to manage different tasks at home?
4. Were you happy with the length/time frame of input from the ACH? Did you anyone ask your preferences?
5. Could you describe for me where your sessions took place, and did you have any preference where your care was delivered?
6. Did you feel you were repeating information to different team members – was there any links with your GP / other healthcare providers – try to draw on examples from their own care.

#### Main Theme: Communication and discharge

1. Could you explain to me your communication with the team, did they provide you with mostly verbal or written information? Did you get all the right information you needed?
2. Did you feel that you were ready to be discharged from the team? How was this communicated to you?
3. How are you managing now at home, did the input from ACH impact your ability to complete daily tasks?
4. Based on your experience, do you have suggestions on how the team could help you better or be developed for other older adults?
5. Has anything been missed from your care – are you waiting for any appointments or has there been any miscommunication.

